# Refining the scope of genetic influences on alcohol misuse through environmental stratification and gene-environment interaction

**DOI:** 10.1101/2024.03.14.24304252

**Authors:** Jeanne E. Savage, Christiaan A. de Leeuw, Josefin Werme, Spit for Science Working Group, Danielle M. Dick, Danielle Posthuma, Sophie van der Sluis

**Affiliations:** Department of Complex Trait Genetics, Centre for Neurogenomics and Cognitive Research, VU University, Amsterdam, The Netherlands; Department of Psychiatry, Robert Wood Johnson Medical School, Rutgers Addiction Research Center, Rutgers University, Piscataway, NJ, USA; Department of Child and Adolescent Psychology and Psychiatry, section Complex Trait Genetics, Amsterdam Neuroscience, VU University Medical Center, Amsterdam, The Netherlands

**Keywords:** genome-wide environment interaction, stratified GWAS, alcohol, trauma, SES

## Abstract

**Background:** Gene-environment interaction (G×E) is likely an important influence shaping individual differences in alcohol misuse (AM), yet it has not been extensively studied in molecular genetic research. In this study, we utilize a series of genome-wide gene-environment interaction (GWEIS) and *in silico* annotation methods with the aim of improving gene identification and biological understanding of AM.

**Methods:** We carried out GWEIS for four AM phenotypes in the large UK Biobank sample (*N* = 360,314), with trauma exposure and socioeconomic status (SES) as moderators of the genetic effects. Exploratory analyses compared stratified GWAS and GWEIS modelling approaches. We applied functional annotation, gene- and gene-set enrichment, and polygenic score analyses to interpret the GWEIS results.

**Results:** GWEIS models showed few genetic variants with significant interaction effects across all gene-environment pairs. Enrichment analyses identified moderation by SES of the genes *NOXA1*, *DLGAP1*, and *UBE2L3,* on drinking quantity and the gene *IFIT1B* on drinking frequency. Except for *DLGAP1*, these genes have not previously been linked to AM. The most robust results (GWEIS interaction *p* = 4.59e-09) were seen for SES moderating the effects of variants linked to immune-related genes on a pattern of drinking with versus without meals.

**Conclusions:** Even in large samples, G×E effects are difficult to detect at the molecular level. Our results highlight several genes and a potential mechanism of immune system functioning behind the moderating effect of SES on the genetic influences on AM. While GWEIS seems to be a preferred approach over stratified GWAS, modelling molecular G×E effects remains a challenge that will require larger consortia and more in-depth phenotypic measurement.

## Introduction

Alcohol misuse (AM) involves multiple dimensions of heavy or problematic alcohol use behaviors that often result in clinical consequences, leading to a major global impact on human health and wellbeing (World Health Organization, 2018). AM has many varied manifestations, but all are strongly influenced by genetics (Dick et al., 2011, Verhulst et al., 2015). However, these genetic influences are subject to moderation by numerous cultural, environmental, and developmental circumstances (Savage et al., 2018b, Young-Wolff et al., 2011, Dick and Kendler, 2012) in a phenomenon known as gene-environment interaction (G×E).

Recent progress in the field through large-scale data collection has begun to reveal the specific genes linked to AM (Kranzler et al., 2019, Liu et al., 2019, Zhou et al., 2020). Such studies typically use a genome-wide association (GWAS) approach to agnostically scan the genome for a statistical association between millions of genetic variants (single nucleotide polymorphisms or SNPs) and a measured phenotype such as quantity of alcohol consumption or alcohol use disorder (AUD) diagnoses. However, much of the ~50% heritability of AM estimated by twin studies (Verhulst et al., 2015) has not yet been accounted for. Explanations for this “missing” heritability include hypotheses that the SNP effect sizes are too small to detect with the current levels of statistical power, that the heritability resides in rare variants that are not easily genotyped with current microarray-based study designs, or that SNP effects are not well characterized because they differ across groups of individuals or environments (i.e., G×E). The field has thus far primarily focused on addressing the first two hypotheses, by increasing the number of participants in GWAS efforts – now frequently including over a million individuals – and expanding whole genome sequencing technologies. However, there have been relatively few attempts to consider other factors like G×E, even though this can readily be investigated with existing data and methodology.

A long history of twin research has demonstrated that genes generally have a stronger influence on AM in contexts in which there are fewer restrictions on alcohol consumption, particularly social restrictions (whether explicit or implicit). For example, the impact of genes is enhanced in countries with low taxes on alcohol sales and in relationships with heavy drinking partners, and decreased in more restrictive contexts such as social groups with a high proportion of drinking abstention and among youth whose access to alcohol is controlled (Dick and Kendler, 2012, Savage et al., 2018b). These moderation effects also appear to differ across the specific dimensions of AM, with, for example, stronger genetic influences on alcohol consumption but weaker genetic influences on AUDs under conditions of high socioeconomic status (SES) (Davis and Slutske, 2018, Barr et al., 2018, Pasman et al., 2020). Strong links between trauma exposure and alcohol in the twin literature (Magnusson et al., 2012) prompted early molecular G×E research which showed that trauma moderates the effects of candidate genes involved in stress response (Baranger et al., 2016, Lieberman et al., 2016) and alcohol metabolism (Sartor et al., 2014). More recently, Polimanti et al. (2018) demonstrated that such moderation effects are detectable in a genome-wide gene-environment interaction (GWEIS) design. A replication study showed the gene-by-trauma effects to be specifically associated with AUD symptoms and not with alcohol consumption (Hawn et al., 2018).

Interaction effects are challenging to detect in molecular genetic studies because their effect sizes are typically an order of magnitude smaller than the already small main effects of SNPs (McClelland and Judd, 1993). However, a GWEIS approach can be fruitful with a sufficiently well-powered sample (Werme et al., 2021, Wendt et al., 2021). Even when individual SNP effect sizes are too small to detect, GWEIS can allow for insight into the underlying biology of a trait through bioinformatics annotation and follow-up methods such as gene-based and gene-set analysis. Approaches using polygenic scores (PGS), which aggregate genetic “risk” across many SNPs, have also demonstrated that molecular G×E functions largely the same way as implied mathematically by twin studies. That is, PGS indexing higher risk for AM tend to show stronger associations in more permissive contexts, such as belonging to deviant peer groups (presenting issues with conduct and antisocial behavior), and weaker effects in more restrictive contexts, such as under high parental involvement (Salvatore et al., 2014, Barr et al., 2017). Like twin-studies, PGS do not reveal the specific genes involved or mechanisms of genetic effect, but the results from such studies echo the notion that individual SNP G×E effects are relevant and waiting to be uncovered.

In an alternative approach to GWEIS, recent studies have shown that stratified GWAS can also improve gene discovery and relevance of genetic associations by splitting the sample into subgroups based on known environmental exposures. For example, a study stratifying groups based on experience of childhood sexual abuse, a major risk factor for depression, uncovered a significant genetic association for depression that was specific to the no-abuse group (Peterson et al., 2017). More generally, studies that subset cases based on known or suspected etiological differences (e.g., early- vs. late-onset Alzheimer’s disease (Kamboh, 2022); typical vs. atypical symptom profiles of depression (Milaneschi et al., 2017); familial vs. complex forms of disorders (Hinney et al., 2010)) can generate meaningful disease insights. This approach takes into account the genetic heterogeneity that is widespread in complex phenotypes, including AM, in which different sets of genetic and/or environmental risk and protective factors are relevant for different individuals. Although statistical power is reduced when stratifying the sample due to the smaller numbers in each subgroup, this is expected to be offset by increased homogeneity, and therefore stronger genetic effects.

To our knowledge, only a few GWEIS and no stratified GWAS have thus far been conducted with AM phenotypes, despite the extensively observed phenomenon of G×E in the twin literature. A stratified GWAS approach fundamentally examines the same question as a modeled G×E approach, but without a formal statistical test comparing whether the genetic effect sizes differ between groups. As both study designs are currently being used in the field, it is important to know which approach is best suited to the aims of gene identification and interpretation for AM. The primary goal of the current study was thus to apply molecular approaches to examine G×E for multiple understudied AM phenotypes in order to uncover genetic variants, genes, and biological pathways that have differential effects on AM given environmental influences. As a secondary exploratory aim, we compared multiple current approaches for implementing G×E on a genome-wide level in order to inform our choice of analysis for the main research question. By incorporating known environmental moderators of the genetic effects on AM, we hope to improve gene discovery and increase the clinical relevance of the identified genetic associations.

## Materials and Methods

### Measures

Data was obtained from the UK Biobank (Bycroft et al., 2018), a population-based sample of adults in the UK with self-report surveys, linked electronic health records, and genotypic data from imputed genome-wide microarrays. The National Research Ethics Service Committee North West–Haydock ethically approved this initiative (reference 11/NW/0382) and participants provided informed written consent. Data were accessed under application #16406.

Genetic analyses in the current study included up to 360,314 unrelated participants of European ancestry with non-missing data. We selected four AM phenotypes for analysis (**Table 1**): a broad indicator of lifetime AUD (BroadAUD), frequency and quantity of alcohol consumption (FreqAlc and QuantAlc), and an indicator of whether alcohol is usually consumed with meals (DrinkMeals). Recent GWAS of these phenotypes (Savage et al., 2023) demonstrated that they represent distinct, clinically relevant dimensions of AM and have robust SNP-based heritability, which is needed to ensure sufficient statistical power for stratified GWAS and G×E analyses. We also selected two robust environmental factors known to moderate the genetic effects on alcohol consumption and problems, respectively: SES, indexed for all participants with the Townsend Deprivation Index (Townsend et al., 1988), and lifetime exposure to traumatic events, measured by self-report in an online survey administered to approximately 157,000 participants (**Table 1**).

**Table 1.** Descriptions of the alcohol phenotypes and environmental measures included in gene-environment interaction analyses.

| Measure | Item name<sup>1</sup> | Description | Field ID<sup>2</sup> |
| --- | --- | --- | --- |
| BroadAUD | broad_aud_ph | Aggregate of lifetime diagnoses of AUD or alcohol-related diseases (alcoholic cirrhosis; Korsakov's syndrome; Wernicke's syndrome; alcoholic ataxia) from medical records, a prescription of an alcohol treatment drug (acamprosate, disulfiram, nalmefene), clinical codes for "personal history of alcoholism", or self-report of an alcohol addiction | 20404,20406 |
| FreqAlc | drinkfreq | Self-report measure of typical drinking frequency | 1558 |
| QuantAlc | quant_ts_gp | Self-report or clinician-reported measures of typical drinking quantity (gm ethanol) per month, log+1 transformed | 1568,1578,1588,1598,1608,4407,4418,4429,4440,4451,4462,5364 |
| DrinkMeals | drink_w_meals | Self-report measure of whether alcohol is typically consumed with or without meals, indexing a broader pattern of drinking habits | 1618 |
| Trauma | - | Lifetime exposure to 10 potentially traumatic events such as assault, abuse, combat experience, a life-threatening accident/illness, or witnessing a violent death | 20488, 20490, 20523, 20524, 20526-20531 |
| SES | - | Socioeconomic status indexed at the time of study enrollment with the Townsend Deprivation Index, a continuous scale score derived from national census measures of material deprivation metrics (e.g., unemployment, home ownership) within a participant's home postcode | 22189 |

*Note:* <sup>1</sup>Item names for alcohol measures relate to the descriptions in Savage et al., 2023 (Supplementary Table 1) which provide greater detail on the derivation of these measures.
<sup>2</sup>Field IDs as listed in the UK Biobank (https://biobank.ctsu.ox.ac.uk/showcase/index.cgi).

## Data Analysis

Based on the twin and epidemiological literature, we chose to investigate five gene-environment pairs: BroadAUD with trauma, QuantAlc with SES, FreqAlc with SES, and DrinkMeals with both trauma and SES. This is because G×E effects have been shown between alcohol problems and trauma and between consumption and SES, as described above, while DrinkMeals is a measure recently shown to be part of a genetically distinct dimension of AM (Savage et al., 2023), but which has not been studied in a G×E context. **Table 2** provides an overview of the analyses performed and the measures involved. The purpose of this pre-selection of environments and phenotypes, as opposed to analyzing all possible combinations, was to reduce multiple testing and avoid data mining.

**Table 2.** Overview of the alcohol phenotypes and environmental measures included in gene-environment interaction analyses.

| Analysis | Outcome phenotype levels | N (N cases / N controls) |
| --- | --- | --- |
| 1. BroadAUD, stratified GWAS, trauma-exposed subset | Case/control | 12,098 / 66,110 |
| 2. BroadAUD, stratified GWAS, non-trauma-exposed subset | Case/control | 4,458 / 43,925 |
| 3. BroadAUD - Trauma, GWEIS-individual | Case/control | 16,534 / 109,887 |
| 4. BroadAUD - Trauma – GWEIS-joint | Case/control | 16,534 / 109,887 |
| 5. FreqAlc – SES, GWEIS-individual | Days per month | 360,314 |
| 6. QuantAlc – SES, GWEIS-individual | Quantity per month | 332,668 |
| 7. DrinkMeals – Trauma, GWEIS-individual | Drink with/not with meals | 51,591/ 16,215 |
| 8. DrinkMeals - SES, GWEIS-individual | Drink with/not with meals | 135,288 / 62,246 |

### G×E model selection

Prior to the primary analyses, we explored multiple ways of examining gene-environment interaction that are often employed in this type of research. Each method seeks to answer the same question (i.e., “are genetic effects equal across levels of an environmental moderator?”) but with different strengths and weaknesses.

1. **Stratified GWAS**: splits the sample into two groups based on a dichotomous environmental exposure and runs a separate GWAS in each group. The results are easily interpretable when there is a significant association in one group but not the other and it is a very simple analysis to carry out. However, there is no formal test o equality across groups to demonstrate whether an effect that is significantly different from zero in one group is also significantly different from the effect in the other group. As such, this is not a true test of G×E and may reflect other influences such as power differences between groups.
2. GWEIS

a. **GWEIS-individual test**: Regression models with an interaction provide a formal test of G×E by means of the regression coefficient and associated *p*-value for the both the main effect term and interaction term, individually. These models are easy to apply in current genetic analysis software but can be difficult to interpret, especially for continuous moderators. There may also be reduced statistical power due to the many covariate-by-SNP and covariate-by-environment interaction terms that need to be included in the model to avoid spurious results (Dick et al., 2015).
b. **GWEIS-joint test**: Regression models with a joint test of significance of the SNP main and interaction effects (rather than only the interaction term) have most of the same strengths and weaknesses as (2a). However, by using a 2 degree of freedom F test with the (usually larger) main effects, they have more statistical power to identify a relevant SNP that may be involved in an interaction, but cannot determine whether identified associations are due to G or G×E specifically (Winham and Biernacka, 2013).

We examined each of these 3 models using BroadAUD and trauma as a test case. For the stratified GWAS (1), the sample was split into subgroups with (n=78,208) or without (n=48,383) a lifetime experience of any type of trauma, and a GWAS of BroadAUD was run within each subgroup. For the GWEIS models (2a & 2b), the trauma measure was treated as a continuous burden score (log+1 transformed to reduce skew) summed across the individual traumatic events. As the aim of this exploratory analysis was to maximize gene discovery, we compared the number of genome-wide significant (*p* < 5e-8) and suggestive (*p* < 5e-6) loci detected between analyses and the strength of associations present. To evaluate the overall concordance between the methods, we estimated genetic correlations using the results from each approach using linkage disequilibrium score regression (LDSC) (Bulik-Sullivan et al., 2015). Genetic correlations were calculated using default software settings and European ancestry LD scores provided with the software. As the GWEIS-joint F-test does not have an associated signed effect direction, we examined Spearman’s correlations between the rank of SNP *p*-values from the GWEIS-joint and each other method. We also carried out gene-set enrichment and polygenic score analyses, described below, to interpret and compare the results from the different models. Here, we aim to see whether adding environmental information can tell us more about the underlying biology of AM and whether convergent results are seen between different models. As there is currently no gold standard among the multiple approaches being used to investigate G×E, we qualitatively compare the gene discovery success and interpretability of results to select the model used for the main analysis of the other AM phenotypes.

### Genetic association analyses

All stratified GWAS and GWEIS analyses were carried out using the generalized linear model (*--glm*) in PLINK2 (Chang et al., 2015) and controlled for sex, genotyping array, and within-ancestry principal components as covariates. Continuous measures (QuantAlc, FreqAlc, SES, and trauma count) were standardized prior to interaction analyses. All interaction models included full SNP×covariate and environment×covariate terms to avoid potential confounding effects (Dick et al., 2015). Full details of the genotyping, quality control, ancestry assignment, and general analysis pipeline have been described previously (Savage et al., 2018a). As G×E analyses can be prone to inflated type I error rates, we applieda sandwich estimator to obtain robust standard errors and adjusted *p*-values for all genome-wide significant loci identified in the main analyses. These analyses were carried out in R version 4.2.2 (R Core Team, 2015) using the method described by Werme et al. (2021) and the *sandwich* package (Zeileis, 2006).

GWAS summary statistics were uploaded to the online platform FUMA (Watanabe et al., 2017) for annotation, gene mapping, and enrichment testing of the associated loci. We used default settings to define associated loci and candidate SNPs based on linkage disequilibrium (LD; *r^2^* > .6) with an independently significant (uncorrected *p* < 5e-8) lead SNP, and then annotated the function of those candidate SNPs from bioinformatics databases. Genes were mapped from candidate SNPs based on their position within the gene, statistical links to expression levels of the gene (eQTLs), or 3-dimensional chromatin interactions between the SNP and the promotor region of the gene. We annotated genes in all significant loci (uncorrected *p* < 5e-8), but consider individual SNPs to be credible only if the robust *p*-value was significant after Bonferroni correction for 5 GWASs (robust *p* < 1e-8).

FUMA was also used to implement gene-based and gene-set testing with MAGMA (de Leeuw et al., 2015). This tests for the enrichment of genome-wide SNP association signal, in aggregate, within individual genes and sets of genes belonging to known biological pathways in the Molecular Signatures Database (curated and gene ontology sets; Liberzon et al., 2011) or with tissue-specific gene expression patterns (GTEx Consortium, 2015). When significant loci were identified, we also used the GENE2FUNC analysis in FUMA to test whether the genes mapped from these loci were similarly enriched in biological pathways and tissues. Per analysis, Bonferroni correction for the number of genes (19,628), gene-sets (10,678), or tissues (54) was applied.

### Polygenic risk score (PRS) prediction

To test replicability of the GWEIS results, we used the summary statistics from these analyses to calculate interaction-based polygenic scores (iPRS^G×E^, as described in Werme et al. (2021)) in an independent sample. For any given SNP and environment, the number of effect alleles each individual carried was multiplied by their observed level of the environmental moderator and by the regression coefficient for the GWEIS SNP×environment interaction. These values were summed across all SNPs to create a single individual-level score, which was then used to predict individual differences in AM measures.

iPRS^G×E^ were calculated in “Spit for Science” (S4S) (Dick et al., 2014), a longitudinal study of college students at a large, urban, public university. The study enrolled multiple cohorts of incoming students who completed yearly self-report surveys and provided a DNA sample. All participants provided informed consent and the S4S study was approved by the university Institutional Review Board. Data was collected and managed by the secure, web-based REDCap system of electronic data capture tools (Harris et al., 2009). The current study used data from *N* = 3,524 S4S participants for whom genotyping and four years of data collection was complete. Samples were restricted to those of European ancestry passing genetic quality control thresholds (Webb et al., 2017, Dick et al., 2014, Peterson et al., 2017). Alcohol use measures assessed in the yearly surveys included questions about typical drinking frequency (number of drinking days per month, averaged across time), typical drinking quantity (gm ethanol per month, following the calculations of (Salvatore et al., 2016) and averaged across time), and *DSM-5* AUD from the Semi-Structured Assessment for the Genetics of Alcoholism (SSAGA; Bucholz et al., 1994). A dichotomous BroadAUD diagnosis was derived based on meeting criteria for any level of AUD (2+ clustered symptoms in the past-year) at any assessment wave. A lifetime count of exposure to traumatic events such as an assault or natural disaster was measured by the Life Events Checklist (Gray et al., 2004). There was no direct measure of SES available, but we used a proxy measure based on the sum of mother’s and father’s years of education.

We derived iPRS^G×E^ in S4S for each of the gene-environment pairs for which GWEIS was applied in UKB, as well as standard PRS from the SNP main effects in the same model. GWEIS summary statistics were weighted using PRS-CS “auto” version and the UKB LD reference panel provided with the software (Ge et al., 2019). Scores were derived in PLINK2 using the –score method. SNPs were first filtered on imputation INFO score > .8, MAF > .05, and missingness < .025. All continuous measures were standardized prior to score creation and analysis. Individual iPRS^G×E^ were used in a linear or logistic regression model in R to predict their corresponding alcohol measures (FreqAlc, QuantAlc, BroadAUD). There was no corresponding measure of DrinkMeals in S4S, so iPRS^G×E^ from the two GWEIS of DrinkMeals were used to predict the other three alcohol measures. All regression models included 10 within-ancestry PCs, sex, and age as covariates, as well as the main effects of the environmental moderator and main effects PRS. Explained variance was calculated as the change in (Nagelkerke’s) *R^2^* between a covariate-only and a covariate + iPRS^G×E^ model.

To facilitate model comparison, we additionally created standard PRS from the stratified trauma/no trauma GWAS of BroadAUD. The procedure described above was used to create and test these PRS except that no environmental measure was included in the score calculation. Stratified PRS were used to predict BroadAUD in subgroups of S4S participants with and without lifetime trauma exposure, respectively. As there were no signed effect sizes produced by the GWEIS-joint model, it was not possible to derive comparative PRS with this model. Bonferroni correction was applied for the 11 models tested.

## Results

### G×E model selection

Exploratory analysis of BroadAUD with trauma indicated that the difference between different model implementations was small, but more significant loci (*p* < 5e-08) were identified in GWEIS than in stratified GWAS (**Figure 1**; 1 vs. 0 loci). The same single locus (chr7: 117544507-117595829, near the gene *CTTNBP2*) was significant in both the GWEIS-individual and GWEIS-joint approaches (**Figure 1c; 1e**). However, the fact that this locus was significant in the GWEIS-individual main effect, while showing no signal in the GWEIS-individual interaction effect (min. *p* = 0.23) suggests that the joint test prioritizes genomic regions that are not necessarily enriched for environmental moderation at even a sub-threshold level. Additionally, several suggestive loci (*p* < 5e-06) with associations specific to the GWEIS-individual interaction were seen (**Figure 1d**), which were not apparent in the GWEIS-joint model.

**Figure 1.**
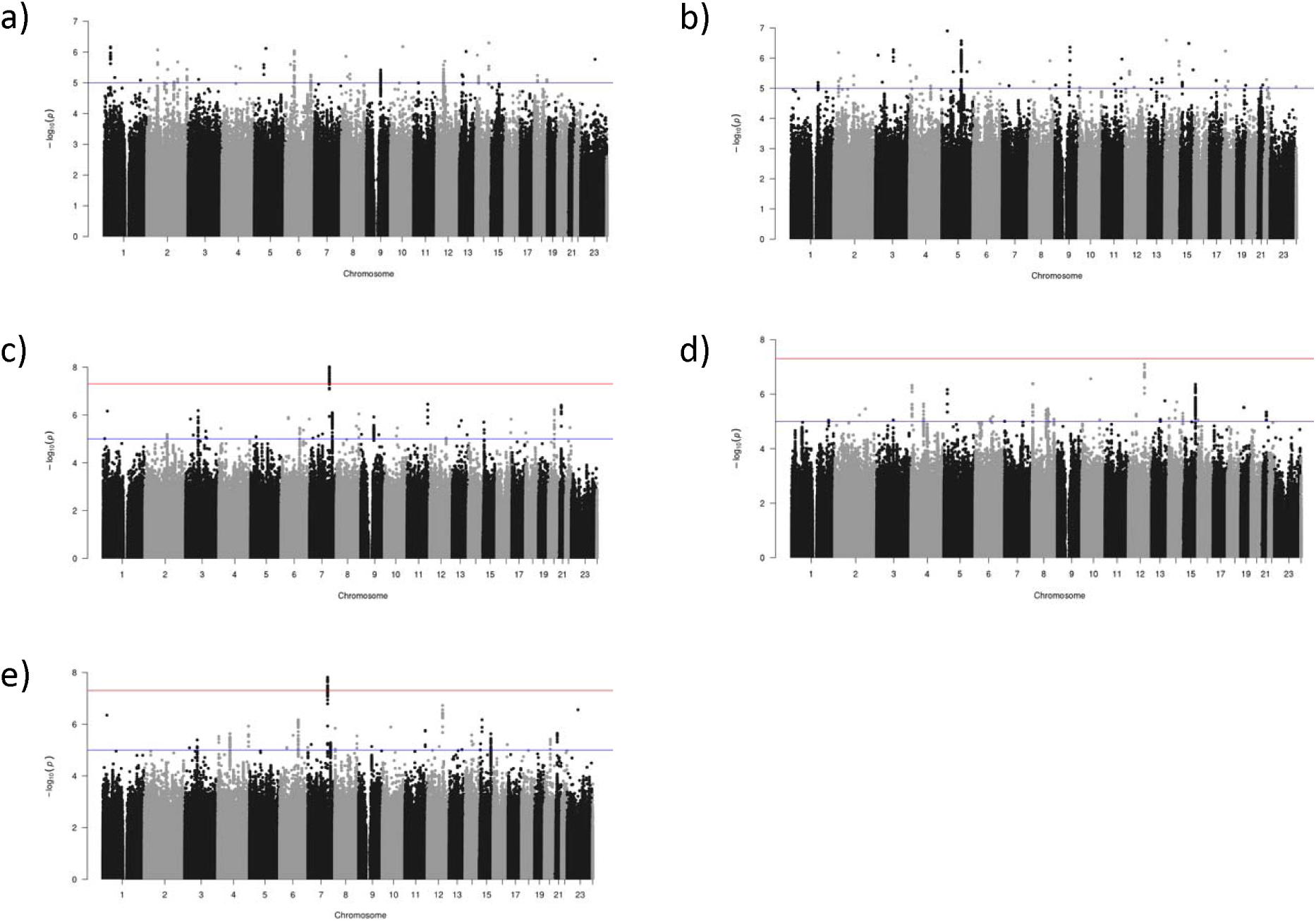
Manhattan plots of the genomic associations for BroadAUD within the a) trauma-exposed and b) non-trauma-exposed subgroups of a stratified GWAS, and for the c) individual SNP main effects and d) individual SNP × trauma interaction effects, and e) joint test of SNP main and interaction effects in a genome-wide gene-environment interaction study (GWEIS).

The mean χ^2^ statistic, indicative of the strength of association signal identified, was similar across methods (stratified trauma group: 1.043, stratified no trauma group: 1.028, GWEIS-individual main effect: 1.016, GWEIS-individual interaction effect: 1.039, GWEIS-joint: 1.038). Genetic correlations between the results of the different modelling approaches are shown in **Table 3**. Genetic effects were largely similar between the stratified groups (*r_g_* = .83) and the stratified GWAS effects were mostly captured by the GWEIS-individual SNP main effects (*r_g_* = .46 - .90). The GWEIS-joint signal largely overlapped with the GWEIS-individual main and interaction effects (*r_g_* = .64 - .65). Gene-set enrichment analysis revealed no significant genes, gene-sets, or tissues for any of the models after Bonferroni correction for multiple testing. Polygenic score analyses showed that, after correcting for 11 models tested (α=.05/11=.0045), neither iPRS^G×E^ from the GWEIS-individual model nor PRS from the stratified GWAS predicted BroadAUD in an independent sample (**Table 4**).

**Table 3.** Genetic correlations between genome-wide association results using multiple modelling approaches for characterizing gene-environment interaction for BroadAUD.

| Model | 1 | 2 | 3 | 4 |
| --- | --- | --- | --- | --- |
| 1. Stratified GWAS - trauma | -- |  |  |  |
| 2. Stratified GWAS – no trauma | 0.83 | -- |  |  |
| 3. GWEIS–individual, main effect | 0.90 | 0.46 | -- |  |
| 4. GWEIS–individual, interaction | -0.07 | -0.59 | -0.38 | -- |
| 5. GWEIS–joint 2df test<sup>a</sup> | 0.12 | 0.27 | 0.65 | 0.65 |

<sup>a</sup> Spearman’s rank-order correlations calculated between SNP p-values as the joint test has no signed effect.

**Table 4.** Polygenic score (PGS) and interaction-based polygenic score-by-environment (iPGS^G×E^) prediction of alcohol phenotypes in an independent validation sample.

| Predictor | Outcome | Main effects PGS |  |  | Interaction iPGS<sup>G×E</sup> |  |  |
| --- | --- | --- | --- | --- | --- | --- | --- |
|  |  | Beta/OR | *P* | *R*<sup>2</sup> | Beta/OR | *P* | *R*<sup>2</sup> |
| BroadAUD stratified (trauma group) | BroadAUD | 0.946 | .138 | .002 | - | - | - |
| BroadAUD stratified (no trauma group) | BroadAUD | 0.895 | .331 | .037 | - | - | - |
| BroadAUD - Trauma | BroadAUD | 0.929 | .048 | .001 | 1.039 | .390 | .000 |
| DrinkMeals - SES | BroadAUD | 0.982 | .655 | .000 | 1.279 | .285 | .000 |
| DrinkMeals - Trauma | BroadAUD | 0.962 | .291 | .000 | 1.066 | .399 | .000 |
| QuantAlc - SES | QuantAlc | 0.027 | .151 | .001 | 0.016 | .557 | .000 |
| DrinkMeals - SES | QuantAlc | 0.003 | .864 | .000 | 0.185 | .087 | .001 |
| DrinkMeals - Trauma | QuantAlc | 0.000 | .998 | .000 | 0.049 | .162 | .001 |
| FreqAlc - SES | FreqAlc | 0.021 | .229 | .000 | 0.027 | .747 | .000 |
| DrinkMeals - SES | FreqAlc | -0.001 | .952 | .000 | 0.058 | .597 | .000 |
| DrinkMeals - Trauma | FreqAlc | -0.006 | .728 | .000 | 0.069 | .052 | .001 |

Overall, comparison of the different approaches did not point to any one model providing substantially greater biological insight than another, but modeled G×E had an edge in gene identification over stratified GWAS. As the GWEIS-individual approach allows for both a statistical test of the interaction and discrimination between the SNP main and interaction effects, we selected this model for all further analyses and refer to it simply as “GWEIS” henceforth.

### Genetic association analyses

For BroadAUD (**Figure 1c-d**), the GWEIS identified no significant loci for the SNP×trauma interaction effect, although one intergenic locus on chromosome 12 (chr12:92736886-92775269) nearly reached genome-wide significance with a *p*-value of 8.02e-08 (robust *p*-value = 4.15e-08). Using positional, eQTL, and chromatin interaction mapping, this locus was linked to 10 genes of which *CLLU1 and CLLU1OS* were the most proximal (**Supplementary Table 1**). There were no significant gene-based or gene-set-based associations.

For QuantAlc, significant SNP×SES interaction effects (**Figure 2a**) were found in loci on chromosome 4 (chr4:24517180-24646772, *p* = 2.96e-08; robust *p* = 2.89e-07), in the gene *DHX15*, and in an intergenic region on chromosome 10 (chr10:44172209-44755269, *p* = 3.28e-08; robust *p* = 6.98e-07). Additionally, two lone significant SNPs were found at chr4:100067606 (*p* = 4.28e-08; robust *p* = 2.31e-07), in a non-coding RNA linked to expression of the alcohol metabolism gene *ADH4*, and on chr5:139695229 (*p* = 4.38e-08; robust *p* = 8.90e-08), in an intergenic region. These four loci were mapped to 17 genes (**Supplementary Table 1**), including *ADH1A, ADH4* and *ADH5*. MAGMA indicated 3 significant gene-based associations (*NOXA1, DLGAP1, UBE2L3*) after Bonferroni correction (**Table 5**), and one significant gene-set association within the GO Biological Pathways set of genes involved in regulation of developmental growth *(p* = 2.31e-06).

**Figure 2.**
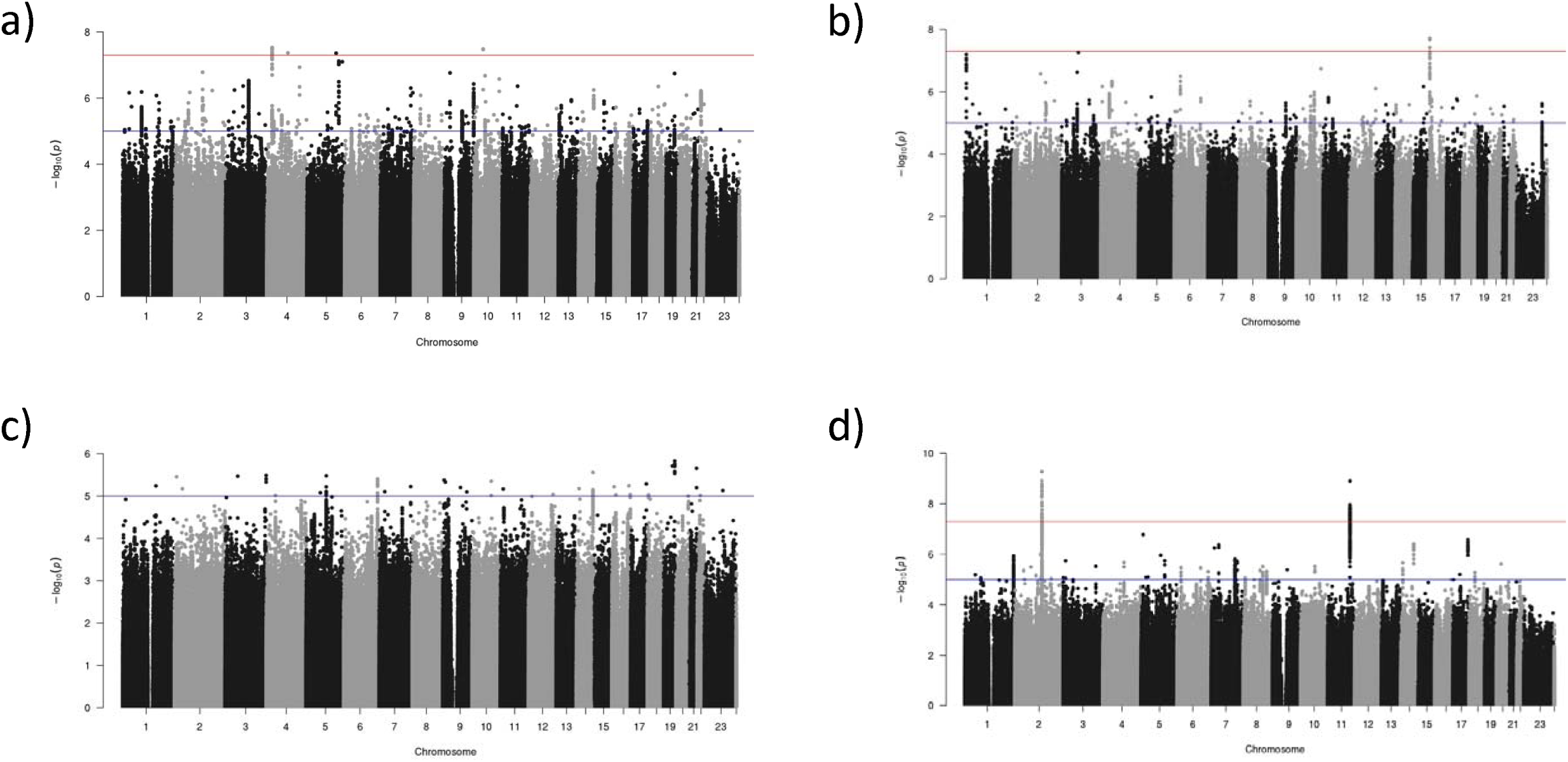
Manhattan plots of the genomic associations for interaction effects from a GWEIS of (a) socioeconomic status (SES) with QuantAlc, (b) SES with FreqAlc, (c) trauma exposure with DrinkMeals, and (d) SES with DrinkMeals.

**Table 5.** Genes with significant enrichment in SNPxSES interaction effects from the GWEIS.

| Analysis | Gene | CHR | Start BP | End BP | # SNPs | Z | P |
| --- | --- | --- | --- | --- | --- | --- | --- |
| QuantAlc-SES | *NOXA1* | 9 | 140317802 | 140328858 | 56 | 4.5643 | 2.51E-06 |
| QuantAlc-SES | *DLGAP1* | 18 | 3496030 | 4455335 | 3480 | 4.583 | 2.29E-06 |
| QuantAlc-SES | *UBE2L3* | 22 | 21903736 | 21978323 | 131 | 4.7598 | 9.69E-07 |
| FreqAlc-SES | *IFIT1B* | 10 | 91137813 | 91144962 | 22 | 5.3841 | 3.64E-08 |
| DrinkMeals-SES | *MAP3K19* | 2 | 135722061 | 135805038 | 272 | 4.7591 | 9.72E-07 |
| DrinkMeals-SES | *R3HDM1* | 2 | 136289025 | 136482840 | 336 | 5.0336 | 2.41E-07 |
| DrinkMeals-SES | *UBXN4* | 2 | 136499189 | 136542625 | 107 | 5.069 | 2.00E-07 |
| DrinkMeals-SES | *LCT* | 2 | 136545410 | 136594750 | 141 | 5.2046 | 9.72E-08 |
| DrinkMeals-SES | *MCM6* | 2 | 136597196 | 136633996 | 88 | 5.2887 | 6.16E-08 |
| DrinkMeals-SES | *DARS* | 2 | 136664247 | 136743670 | 152 | 4.5928 | 2.19E-06 |
| DrinkMeals-SES | *SIK2* | 11 | 111473115 | 111601577 | 339 | 4.9598 | 3.53E-07 |
| DrinkMeals-SES | *PPP2R1B* | 11 | 111597632 | 111637151 | 99 | 5.0392 | 2.34E-07 |
| DrinkMeals-SES | *ALG9* | 11 | 111657010 | 111750149 | 224 | 5.5454 | 1.47E-08 |
| DrinkMeals-SES | *FDXACB1* | 11 | 111744780 | 111751967 | 21 | 5.3291 | 4.94E-08 |
| DrinkMeals-SES | *CRYAB* | 11 | 111779289 | 111794446 | 33 | 4.9116 | 4.52E-07 |
| DrinkMeals-SES | *HSPB2-C11orf52* | 11 | 111783460 | 111797595 | 30 | 4.8833 | 5.22E-07 |
| DrinkMeals-SES | *C11orf52* | 11 | 111788756 | 111797596 | 18 | 5.0699 | 1.99E-07 |
| DrinkMeals-SES | *DIXDC1* | 11 | 111797868 | 111893308 | 195 | 5.37 | 3.94E-08 |
| DrinkMeals-SES | *DLAT* | 11 | 111895538 | 111935114 | 82 | 5.2919 | 6.05E-08 |
| DrinkMeals-SES | *PIH1D2* | 11 | 111934734 | 111944998 | 22 | 5.524 | 1.66E-08 |
| DrinkMeals-SES | *C11orf57* | 11 | 111944810 | 111955874 | 16 | 5.3897 | 3.53E-08 |
| DrinkMeals-SES | *TIMM8B* | 11 | 111955524 | 111957522 | 2 | 5.0433 | 2.29E-07 |
| DrinkMeals-SES | *SDHD* | 11 | 111957497 | 111990353 | 66 | 5.1472 | 1.32E-07 |

Note: genes shown are significant after Bonferroni correction for 19,628 protein-coding genes ( $p < .05/19628 = 2.55e-06$ )

For FreqAlc (**Figure 2b**), GWEIS identified one significant locus for the SNP×SES interaction (chr16:5143432-5176998, *p* = 1.91e-08; robust *p* = 5.73e-08). The genes *ALG1* and *FAM86A* were implicated by positional mapping, including a candidate exonic nonsynonymous variant in *FAM86A*, and the genes *MGRN1* and *SEC14L5* were linked via eQTL SNPs in the locus which affect expression levels of these genes in the brain (**Supplementary Table 1**). The gene *IFIT1B* was identified in gene-based analysis (*p* = 4.50e-08). There were no gene-sets or tissues enriched for association.

For DrinkMeals (**Figure 2c**), there were no significant SNP interaction effects in the model with trauma count as a moderator. With SES as a moderator (**Figure 2d**), there were two loci with significant SNP×SES interactions: chr2:135771974-136787402 (*p* = 5.20e-10; robust *p* = 3.35e-03) and chr11:111541124-111989063 (*p* = 1.23e-09; robust *p* = 4.59e-09; considered credible after Bonferroni correction). Gene-based tests implicated 19 genes (**Table 4**), several of which contained exonic nonsynonymous variants among the candidate SNPs: *LCT* (chr2), *FDXACB1, ALG9, DLAT, PIH1D2,* and *SDHD* (chr 11). The 94 genes mapped by these loci (**Supplementary Table 1**) were enriched for expression in breast, spleen, visceral adipose, and esophagus tissues (*p* < 4.36e-04). They were also enriched in multiple gene-sets involved in immune system functioning, particularly in the immunological signatures of plasmacytoid dendritic cells (**Table 6**).

**Table 6.** Gene-sets with significant enrichment in genes mapped by the GWEIS of SNPxSES interaction effects predicting DrinkMeals.

| GeneSet | N genes | n overlap | P-value |
| --- | --- | --- | --- |
| GSE29618_MONOCYTE_VS_PDC_DN | 187 | 27 | 1.20e-32 |
| GSE29618_PDC_VS_MDC_UP | 189 | 27 | 1.62e-32 |
| GSE29618_PDC_VS_MDC_DAY7_FLU_VACCINE_UP | 190 | 26 | 8.62e-31 |
| GSE29618_BCELL_VS_PDC_DN | 195 | 24 | 2.89e-27 |
| GSE29618_BCELL_VS_PDC_DAY7_FLU_VACCINE_DN | 195 | 24 | 2.89e-27 |
| GSE29618_MONOCYTE_VS_PDC_DAY7_FLU_VACCINE_DN | 192 | 23 | 7.44e-26 |
| GSE17301_ACD3_ACD28_VS_ACD3_ACD28_AND_IFNA5_STIM_CD8_TCELL_DN | 193 | 11 | 2.74e-09 |
| GSE16385_ROSIGLITAZONE_IL4_VS_IL4_ALONE_STIM_MACROPHAGE_12H_UP | 193 | 9 | 4.54e-07 |
| GSE29618_PRE_VS_DAY7_FLU_VACCINE_MDC_UP | 188 | 8 | 4.04e-06 |

*Note: gene-sets shown are significant after Bonferroni correction for 10,678 pathways in the Molecular Signatures Database curated and gene ontology sets ( $p < .05/10678=4.68e-06$ )*

### PRS prediction

Across all alcohol phenotypes, PRS and iPRS^G×E^ based on the GWEIS results did not significantly predict corresponding alcohol measures in an independent sample (**Table 4**).

## Discussion

### Summary

In this study, we conducted a large-scale GWEIS on multiple dimensions of alcohol misuse in a population-based sample. By concentrating on phenotypes with previously demonstrated SNP heritability and environmental factors (trauma and SES) with known moderating effects on AM, we maximized our *a priori* likelihood of identifying AM-relevant gene-environment interactions. Despite this, we found few genomic regions with strong evidence for G×E and poor out-of-sample prediction from interaction-based polygenic risk scores. There was only one locus with convincing evidence for G×E at an individual SNP level (i.e., robust *p*-value meeting the Bonferroni-corrected genome-wide significance threshold of 1e-8). This was found for SES moderating the effect of SNPs mapped to immune-related genes on a pattern of drinking with vs. without meals. Our aggregate gene-based results, however, nominate several candidate genes which seem to interact with SES to influence alcohol consumption and drinking patterns.

### Interpretation of G×E effects

Trauma exposure has long been viewed as a catalyst for the manifestation of genetic risk for AM (Magnusson et al., 2012). Yet, in this study, we found little evidence that the effects of genetic variants are moderated by trauma or that they behave differently in individuals with vs. without trauma exposure. This is in contrast to previous GWEIS in smaller samples (*N*=20,000-50,000 versus the current *N*=120,000-360,000) which demonstrated several significant SNP×trauma interactions for AM (Cheng et al., 2021, Polimanti et al., 2018). This lack of replication may be due to the more specific focus of these other studies on childhood trauma exposure (Cheng et al., 2021) or in a mostly military sample (Polimanti et al., 2018). Genetic heterogeneity is a major challenge in gene identification for AM (Wong and Schumann, 2008), and variability in the type, timing, or context of trauma exposure may add an additional layer of complexity obscuring the underlying genetic influences. However, it may also be that earlier results were false positives which fail to replicate in larger studies, as has been the case for much G×E research.

For interactions involving SES, we are cautious of interpreting individual SNP effects as most (robust) *p*-values did not survive multiple testing corrections. However, the gene-based tests, which aggregate individual SNP G×E effects, pointed to several promising candidate genes. For drinking quantity, these included the NADPH oxidase activator *NOXA1*, DLG associated protein *DLGAP1*, and ubiquitin conjugating enzyme *UBE2L3,* and, for frequency, the interferon induced protein with tetratricopeptide repeats *IFIT1B*. Except for *DLGAP1*, these genes have not previously been linked to AM. The GWASatlas (Watanabe et al., 2019) reports that *NOXA1* is associated with metabolic phenotypes like cholesterol and body fat, while *UBE2L3* has strong reported associations with numerous immunological traits/diseases and *IFIT1B* has been linked to smoking. *DLGAP1* has associations across a broad range of domains of health and behavior including an early GWAS of alcohol dependence (Wang et al., 2013). Genes whose effects on drinking quantity were moderated by SES were also enriched in regulation of developmental growth, suggesting that growth and/or metabolic processes related to drinking are broadly impacted by SES. Most interesting were the results for DrinkMeals, an AM phenotype which has recently been shown to represent an important distinct dimension of AM (Savage et al., 2023). G×E enrichment in multiple genes related to immune function suggests that genetic predispositions for immune-related conditions might differentially impact patterns of alcohol use at high versus low SES. For example, this could indicate differential patterns - influenced by income or other SES-related factors - of avoiding alcohol versus self-medicating in response to health problems.

To our knowledge, there are no other GWEIS examining SES with which we can compare these results, so the mechanism of action behind these interactions remains to be investigated. PRS prediction into a sample of college students did not show evidence of replication. It is uncertain whether this is due to the results being unreliable, the effect sizes too small, or the AM phenotypes and environmental moderators being too different across the discovery and validation samples (genetic heterogeneity). As substantial sub-threshold association signal was evident in the GWEIS, it may be that even larger samples and/or more refined measures are needed to obtain sufficient statistical power.

### Strategies for modelling G×E

Through comparison of multiple G×E study designs, we also suggest that modelling main and interaction effects in a GWEIS individually is a more informative approach for characterizing molecular G×E, than stratified GWAS or a GWEIS with a joint test of the SNP main and interaction effects. We did not find substantial differences between these models in terms of their gene identification success or predictive power, but we found the GWEIS individual test model to be the most interpretable while formally comparing the difference in SNP effect sizes as a function of the level of the environmental moderator. However, our results may be limited by the power of the phenotype these models were applied to. A simulation study that systematically compares these models under different conditions could help to guide future G×E research.

### Limitations and conclusion

Despite substantial increases in sample size and phenotypic breadth over previous GWEIS of AM, the results indicate that we still have limited power to detect G×E at the individual SNP level, although the twin literature indicates that they must exist. Until this issue is resolved, the biological insight provided by such findings is necessarily low. Our study is limited by the relatively shallow measures collected and the known biases in drinking behavior (Xue et al., 2021) due to the “healthy volunteer” bias in UKB (Fry et al., 2017). Nonetheless, we have highlighted several genes and a potential mechanism of immune system functioning behind the moderating effect of SES on the genetic influences on AM. We also emphasize the importance of considering specific dimensions of AM, as there was little overlap in the results between AUD, consumption, and drinking patterns. We conclude that GWEIS may be a preferred approach over stratified GWAS but modelling molecular G×E effects remains a challenge. Larger consortia, more in-depth measures, and longitudinal data collection are needed to resolve the complex gene-environment interplay underlying AM.

## Acknowledgements

This research was funded by a grant to J.E.S. (VI.VENI.201G-064) from The Netherlands Organization for Scientific Research (NWO) and has been conducted using the UK Biobank Resource (application no. 16406). We would like to thank the many UK Biobank participants and staff. Analyses were carried out on the Genetic Cluster Computer hosted by the Dutch National computing and Networking Services SURFsara. Spit for Science has been supported by Virginia Commonwealth University, P20AA017828, R37AA011408, K02AA018755, P50AA022537, and K01AA024152 from the National Institute on Alcohol Abuse and Alcoholism, and UL1RR031990 from the National Center for Research Resources and National Institutes of Health Roadmap for Medical Research. This research was also supported by the Center for the Study of Tobacco Products at Virginia Commonwealth University. The content is solely the responsibility of the authors and does not necessarily represent the views of the funding agencies. The funding agencies had no role in the study design, data analysis, manuscript preparation, or decision to submit for publication. We would like to thank the Spit for Science participants for making this study a success, as well as the many University faculty, students, and staff who contributed to the design and implementation of the project.

## Data availability

Genome-wide summary statistics will be made publicly available via https://ctg.cncr.nl/software/summary_statistics/ upon publication. Raw data from this study are available to qualified researchers via UK Biobank (https://www.ukbiobank.ac.uk/) and dbGaP (phs001754.v4.p2).

